# Combined [¹¹C]PBR28 PET and Quantitative T1 Mapping Reveal Choroid Plexus Alterations in Multiple Sclerosis

**DOI:** 10.1101/2025.11.10.25339910

**Authors:** Sneha Senthil, Risavarshni Thevakumaran, Stephan Blinder, Alexey Kostikov, Rozie Arnaoutelis, Pedro Rosa-Neto, G.R. Wayne Moore, Douglas Arnold, Sridar Narayanan, David A. Rudko

## Abstract

**Background:** The choroid plexus (CP), central to cerebrospinal fluid (CSF) regulation and immune cell trafficking, has been implicated in multiple sclerosis (MS).

**Objectives:** We investigated TSPO-PET standardized uptake value ratio (SUVR) in the CP, with relation to T□ relaxation times and CP volume using 7T MRI.

**Methods:** Quantitative T1 (qT_1_) maps were obtained from 38 MS participants and 21 healthy controls (HCs) using 7T MRI. Within 4 weeks, PET imaging was conducted with [¹¹C]PBR28 to measure SUVR in the CP. Group differences and associations in CP SUVR, qT_1_, and CP volume were assessed.

**Results:** CP SUVR, CP qT_1_, and head□size-corrected CP volumes were higher in MS than HCs (*p<0.05*). CP volume associated strongly with CP qT_1_ (*r=0.52*, *p<0.05*). In MS, CP SUVR inversely correlated with qT_1_ (ρ= –0.45, *p*<0.05). No significant associations were observed with disability scores, white□matter lesion burden and cortical lesion counts.

**Conclusions:** Our findings support elevated microglial/macrophage activation in CP alongside structural alterations. CP volume emerges as a complementary MRI marker of inflammation rather than a direct surrogate for TSPO-PET tracer uptake. The inverse CP SUVR–qT□ relationship in MS suggests that chronic extracellular expansion can accelerate tracer washout.

## Introduction

Multiple sclerosis (MS) is the most prevalent inflammatory demyelinating disease of the central nervous system (CNS), characterized by immune-mediated injury to myelin and axons. The most common clinical form of MS is relapsing-remitting MS (RRMS), in which disease exacerbations are followed by periods of relative inactivity and recovery, although there is substantial variability in clinical course among people with MS (pwMS). It has been shown recently that most people with RRMS experience disability worsening that occurs independently of relapses (1). This is referred to as PIRA (progression independent of relapse activity), a phenomenon that became observable due to the advent of highly-effective disease-modifying drugs that suppressed most relapses (2, 3). Clinically, sustained PIRA contributes to the transition from relapsing to secondary progressive MS (SPMS).

In recent years, there has been a growing interest in the role of the choroid plexus (CP) in MS pathogenesis as it has been proposed as a key site for immune cell migration into the CNS (4, 5). The CP, a highly-vascularized structure within the brain ventricles, is responsible for producing cerebrospinal fluid (CSF) and maintaining homeostasis of the blood–CSF barrier (BCSFB), by regulating the entry of immune cells and solutes into the CNS (6). Disruption of the BCSFB can facilitate immune cell entry, edema formation, and altered CSF composition-processes that are intimately linked with MS disease activity (7).

Post□mortem studies in progressive MS have revealed the CP is frequently infiltrated by leukocytes and exhibits fibrinogen deposition, with CP inflammation correlating with perivascular inflammation and the burden of active demyelinating lesions (8). Advanced *in vivo* MRI studies corroborate these pathological observations and have demonstrated both structural and functional alterations of the CP in MS. Additionally, volumetric analyses have demonstrated that CP enlargement may discriminate MS from its mimics, including neuromyelitis optica spectrum disorders (9). CP enlargement has also been linked with measures of periventricular demyelination (10-12). Interestingly, dynamic contrast□enhanced (DCE) MRI studies have evaluated abnormally elevated CP permeability, with higher values of plasma volume and efflux rate of gadolinium contrast from blood plasma into the tissue extravascular extracellular space (K_trans_) in MS compared to controls, even during clinically latent phases (13). Further, quantitative MRI (qMRI) using T_1_ mapping has shown that the degree of CP enhancement strongly correlates with markers of diffuse brain tissue injury (14). Furthermore, positron emission tomography (PET) studies report ∼18–20% higher uptake of tracers sensitive to translocator protein (TSPO) expression in MS CP versus controls, particularly in relapsing–remitting cases (10), indicating heightened innate immune activation driven by activated microglia and infiltrating macrophages. Nonetheless, it remains unclear whether elevated TSPO-PET standardized uptake value ratios (SUVR) in CP, which primarily reflects microglial/macrophage activation, is related to chronic BCSFB breakdown and whether the interaction between these processes may perpetuate chronic neuroinflammation and ultimately contribute to neurodegeneration processes in MS.

In this multimodal imaging study, we leveraged quantitative T_1_-based relaxometry derived from ultra-high-field (7T) MRI and high-resolution [¹¹C]PBR28 PET to interrogate alterations in CP pathology in MS. Our primary objectives were to determine whether (1) CP TSPO-PET SUVR is elevated in pwMS compared to healthy controls, (2) qT□ of the CP is prolonged in MS, (3) qT_1_ and SUVR of the CP are interrelated and (4) CP volume is related to TSPO tracer uptake. We additionally evaluated whether CP SUVR and CP qT□ were associated with clinical disability (EDSS), CP morphology (head-size corrected CP volume), and lesion burden (white matter lesion, WML, volume, intracortical and leukocortical lesion counts).

## Methods

### Participant Recruitment

Thirty□eight pwMS (19 relapsing–remitting, 19 progressive; age 55.45 ± 10.97 years; 12 male, 26 female; EDSS 3.84 ± 1.85; disease duration 24 ± 13.5 years), followed at the MS Clinic of the Montreal Neurological Institute-Hospital, and twenty□one age□ and sex□matched healthy controls (age 51.05 ± 12.62 years; 9 male, 12 female) were recruited. MS participants met the 2017 revised McDonald criteria (15), were ≥ 18 years old, and had been untreated or on stable disease□modifying therapy (DMT) for at least six months. Exclusion criteria included the existence of other neurological disorders, relapse or steroid treatment within 30 days, claustrophobia, and MRI contraindications (e.g., metallic implants, pacemakers). Healthy controls had no history of neurological or psychiatric illness, no prior brain injury or neuroinflammatory disease, and no contraindications to MRI. Detailed demographic information characterizing the cohort is presented in Table 1. All study procedures were approved by the Research Ethics Board of the Montreal Neurological Institute-Hospital. Written informed consent was obtained from all study participants.

**Table 1:** Detailed demographic information of the recruited participant cohort.

| **Characteristics** | **Full MS cohort (n = 38)** | **RRMS (n = 19, 50%)** | **PMS (n = 19, 50%)** | **HC cohort (n=21)** |
| --- | --- | --- | --- | --- |
| Age in years [Mean(SD)] | 55.4(11.0) | 49.4(9.8) | 61.5(8.6) | 51.0 (12.6) |
| Female (%) | 26(68.4%) | 12(63.2%) | 14(73.7%) | 12(57.1%) |
| Binding affinity:<br>HAB;MAB;LAB | 18;14;6 | 12;5;2 | 6;9;4 | 10;10;1 |
| EDSS [median(IQR)] | 4.2(3.3) | 2.5 (1.8) | 6.0 (2.5) | NA |
| Recent relapse activity (in the last one year) | 7(18.4%) | 7(36.8%) | 0 | NA |
| Average WML volume (mm<sup>3</sup>)<br>[Mean(SD)] | 17740.2(21738.8) | 7503.1(8986.9) | 30339.7(26268) | NA |
| Total Number of Gd-enhancing lesions | 0 | 0 | 0 | NA |
| Leukocortical lesion count<br>[Median(IQR)] | 2.5(1.5) | 1(2) | 4(3) | NA |
| Intracortical lesion count<br>[Median(IQR)] | 0(0.5) | 0(0.5) | 0(0.5) | NA |
| Disease duration (years)<br>[Mean(SD)] | 24(13.5) | 30.0(15.5) | 18.0(7.5) | NA |
| On DMT (None, Low-Medium efficacy, High<br>efficacy) | 20,8,10 | 5,7,7 | 15,1,3 | NA |

WML = white matter lesion, DMT = Disease modifying treatment, SD = standard deviation, IQR=interquartile range ,min=minimum, max=maximum, RRMS = relapsing-remitting multiple sclerosis, PMS = progressive multiple sclerosis.

### MRI

#### MRI acquisition

All participants were scanned using a 7T whole-body MR scanner (MAGNETOM Terra, Siemens Healthineers, Erlangen, Germany) with an 8 Tx/32 Rx-channel head coil (Nova Medical Inc., Wakefield, MA, USA) in parallel transmission (pTx) mode. The imaging protocol included an MP2RAGE acquisition, with the following sequence parameters: isotropic resolution = 0.7mm^3^, FOV = 240×240×172 mm^3^, TI1/TI2 = 800 ms/2700 ms, TR/TE = 6000 ms/2.74 ms, acquisition time = 10:14 min. A T1 map was computed, based on the MP2RAGE acquisition, using the Siemens MapIt software. A 3D T2-weighted Fluid Attenuated Inversion Recovery (FLAIR) sequence was also acquired in the MS cohort for lesion segmentation, with the following sequence parameters: isotropic resolution = 0.7mm^3^, FOV = 225×225×179.2 mm^3^, TR/TE = 10560 ms/301 ms, TI = 2500 ms, acquisition time= 9:32. All scans were visually inspected to confirm the absence of significant motion artifacts that could compromise subsequent image analysis.

#### MRI post-processing and analysis

MR images were processed using the MINC toolkit (https://bic-mni.github.io/) (16). Specifically, images for each MS participant were individually co-registered to the native MP2RAGE space to ensure spatial alignment. A brain mask was then generated based on the MP2RAGE uniform intensity (UNI), denoised anatomical image using a U-net-based deep learning segmentation method (17). The brain mask was resampled and applied to the MP2RAGE-derived and FLAIR images collected in the study. Intensity inhomogeneities in the 7T FLAIR images were corrected using the N4 bias field algorithm (18). Mean qT_1_ of CP and volume measures were also collected by applying each subject’s CP mask to their respective qT_1_ map.

#### Choroid Plexus segmentation

CP segmentations for each participant were manually performed using the MP2RAGE images. While the denoised MP2RAGE uniform (UNI) image derived from the Siemens MapIt software provides excellent whole-brain, T1-weighted contrast, it often renders the ventricular CSF signal dark. This makes the CP appear less distinct against the surrounding CSF. By contrast, the second inversion time image (INV2 image) offers superior visibility of the CP due to its brighter CSF signal and better CP to CSF contrast. We observed that using the INV2 image enhanced the delineation of CP boundaries by providing greater separation between CP tissue and adjacent ventricular spaces, facilitating more accurate segmentation (Figure 1). The MP2RAGE-derived INV2 image was, therefore, used as the primary reference for CP identification, with additional confirmation using the FLAIR images, when available.

**Figure 1:**
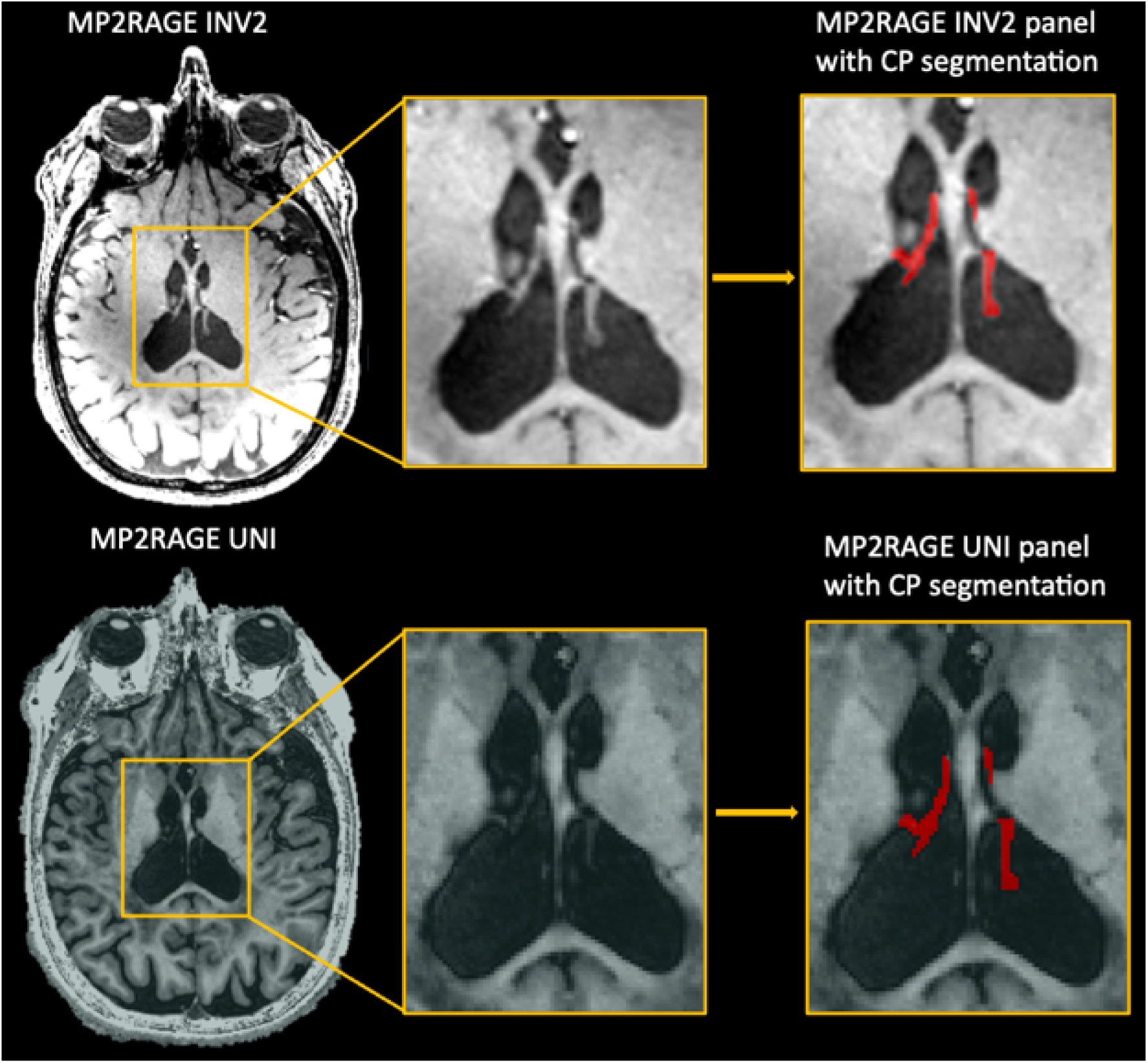
MP2RAGE INV2 and UNI axial images from a representative PMS participant (top row and bottom row, respectively). Panels with CP segmentation of each image are shown on the right. The UNI image is windowed in order to delineate the CP mask. The INV2 image demonstrates markedly superior CP-to-surrounding tissue contrast, highlighting its advantage for accurate CP segmentation.

#### Lesion segmentation

White-matter lesion (WML) segmentation was performed as previously described in Thevakumaran et al. (19). As well, intracortical and leukocortical lesions were likewise carefully, manually segmented using ITKSNAP based on T1- and T2-weighted FLAIR contrasts (≥ 3 contiguous slices, ≥ 4 voxels/slice) (19). WML volume for each MS participant was calculated and both intracortical and leukocortical lesion counts were taken into account during further analyses.

### PET Imaging

#### PET acquisition

In addition to a 7T MRI scan, all participants also underwent a 90-minute [^11^C]PBR28 PET scan on a high resolution research tomograph (ECAT HRRT, Siemens, Knoxville, TN, USA) PET scanner (20). PET emission data was acquired in list mode, following an intravenous bolus injection of a [^11^C]PBR28 radiotracer produced in-house (mean ± SD dose of 10.3 ± 0.5 mCi in MS patients; 10.6 ± 0.9 mCi in healthy controls, *P =* 0.2 by unpaired t-test). Data was then reconstructed using the 3D OP-OSEM algorithm with a reconstructed voxel size of 1.22mm^3^ (20). Participants were genotyped for the Ala147Thr polymorphism in the TSPO gene to determine binding affinity of [^11^C]PBR28 to TSPO. A supervised clustering algorithm (SVCA) was then applied to each PET time–activity curve to extract a subject-specific pseudo□reference region for standard uptake value normalization (21). Normalizing SUV to such a reference region yielded SUVR values that more accurately reflected specific [^11^C]PBR28 binding to TSPO, independent of inter-individual variability of radiotracer delivery, molar activity, and nonspecific binding (21). High affinity binders (HABs) and medium affinity binders (MABs) were considered. PET data from low□affinity binders (LABs) were excluded from further analysis due to negligible specific signal in the CP, which could result in unreliable quantification (22).

#### PET data processing

PET data were reconstructed using the 3D ordered-subsets expectation maximization (OP-OSEM) algorithm, applying all standard corrections and motion correction, on a 256 × 256 × 207 grid with a reconstructed voxel size of 1.22 mm³ (23). PET emission frames were first corrected for head motion using frame-to-frame rigid registration and then summed to generate a static 60–90□min post□injection image. All frames and the static image were coregistered to the participant’s 7□T MP2RAGE UNI□DEN volume using mutual information□driven registration (24).

Next, interregional partial□volume effects (PVEs) were corrected for each dynamic frame and the static PET image using the PETPVC toolbox (version 1.2.0-b, https://github.com/UCL/PETPVC) (25). The combined Labbé and region□based voxel□wise (Labbé□RVC) PVC approach was applied using a 3□mm FWHM isotropic Gaussian approximation of the scanner point□spread function (PSF) (26, 27), with anatomical tissue masks (ventricles, CP, cortical GM, WM) derived from the MP2RAGE segmentations.

Following PVC, voxel-wise standardized uptake values (SUVs) were computed by normalizing the decay-corrected static PET activity concentrations to each subject’s injected dose and body weight. To derive SUVR maps, each SUV in each region of interest was divided by the mean SUV within the SVCA-derived pseudo-reference region in cortical gray matter. The CP segmentation mask was then applied to the SUVR map to obtain measures of TSPO-specific tracer uptake in the CP.

### Statistical Analysis

All statistical analyses were performed using R (v4.4.0; R Core Team, 2024). Group differences in CP SUVR, qT□ of CP, and head-size normalised CP volume, between MS and controls were assessed using unpaired one□tailed *t*-tests, assuming higher mean values in the MS cohort. When normality (Shapiro–Wilk) was violated, one-tailed Wilcoxon rank-sum tests were used. An exploratory two-tailed *t*-test was then performed in the RRMS subgroup. Spearman partial correlation was calculated to assess the relationship between mean qT_1_ of the CP and CP TSPO-PET SUVR in HC and MS subjects separately.

Linear regression models were applied to examine the association between CP SUVR and CP qT□ with the native CP volume. Model fits were evaluated and nested models were compared using an extra (partial) sum-of-squares F-test based on the reduction in residual sum of squares. Spearman’s correlation was used to test associations of CP SUVR and CP qT_1_ with EDSS score, CP volume and lesion metrics (including WML volume, intracortical and leukocortical lesion counts). Age, sex, CP volume, and TSPO binding affinity (HAB vs. MAB) were included as covariates of no interest (nuisance variables) when found to be significant using Akaike information criterion (AIC) and model comparison F-tests. Secondary analyses were Bonferroni-corrected to control for multiple comparisons. Statistical significance was defined as *p*□<□0.05.

## Results

### Characteristics of study population

Detailed demographic and clinical characteristics of the recruited cohort are shown in Table 1. PwMS comprised equal proportions of RRMS (50%) and PMS (50%) participants. 52.6% of pwMS participants were on Disease Modifying Therapies (DMTs) at the time of their participation. When stratified based on drug efficacy, 26.3% of pwMS were on low-medium efficacy drugs and 73.6% were on high efficacy drugs. 18.4% of participants had at least one relapse in the one year preceding their visit and all of these were of the RRMS subtype. Visual assessment of post-contrast FLAIR and T1-weighted images revealed no Gd-enhancing lesions. A total of 26.3% participants were identified as low-affinity binders (LABs) based on TSPO genotyping and were excluded from subsequent PET analyses.

### Group comparisons of choroid plexus qT_1_, SUVR and volume

Table 2 summarizes the CP data derived from qT1 maps and SUVR maps for each participant group. Figure 2 presents a qT1 map and co-registered SUVR map for a representative RRMS subject with associated CP segmentations. Compared to HCs, pwMS showed significantly higher CP SUVR values, indicating elevated [¹¹C]PBR28 uptake (p<0.05; Figure 3A). Exploratory subgroup analyses were performed within the MS cohort to identify drivers of elevated CP SUVR. The RRMS subgroup appeared to drive the overall increase in CP SUVR (p<0.05).

**Figure 2:**
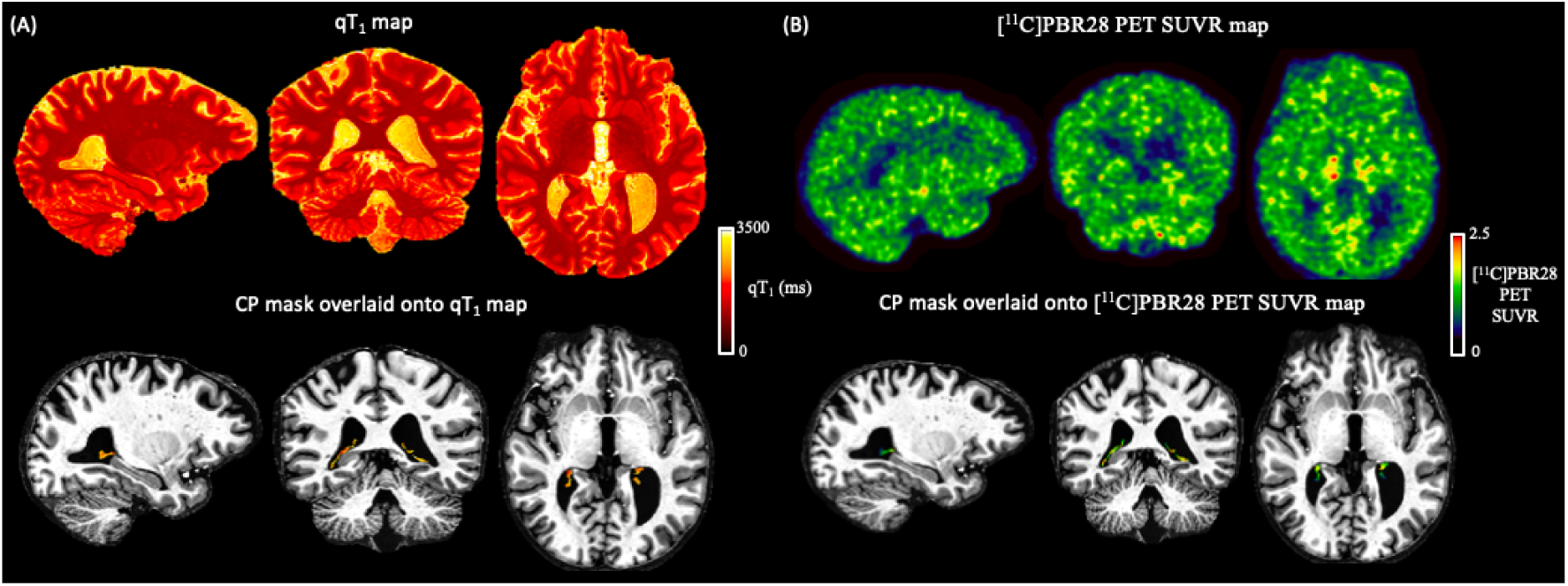
Representative sagittal, coronal, and axial views of qT_₁_ and [¹¹C]PBR28 SUVR maps of the CP in a participant with RRMS. (A) qT_₁_ maps showing regional variations within the CP (top: native qT_₁_ map; bottom: qT_₁_ map within the CP mask overlaid on the MP2RAGE UNI T1-weighted image). (B) Corresponding [¹¹C]PBR28 SUVR maps from the same participant (top: SUVR map; bottom: SUVR map within the CP mask overlaid on the MP2RAGE UNI T1-weighted image), highlighting regional heterogeneity of TSPO binding within the CP.

**Figure 3:**
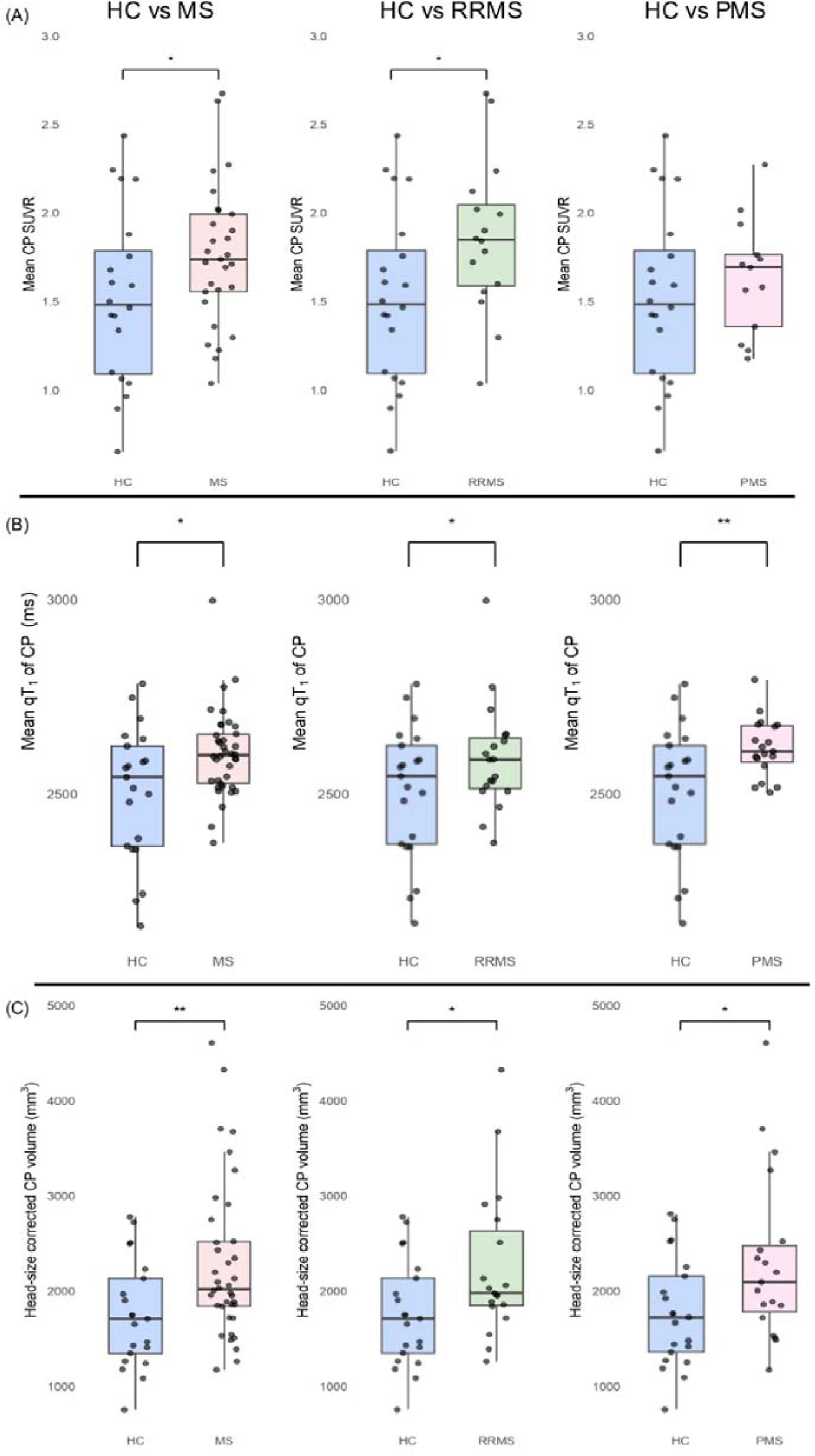
Box plots showing group differences in (A) mean CP [¹¹C]PBR28 SUVRs, (B) mean qT_1_ of CP (in ms) and (C) Head-size corrected CP volume (in mm^3^). Plots compare the HC group with MS, RRMS, and PMS. Each box represents the interquartile range with a median line; whiskers extend to 1.5x the interquartile range. Individual points represent participants. Significance markers (*p < 0.05, **p < 0.01) indicate one-tailed t-test results (MS > HC).

**Table 2:**
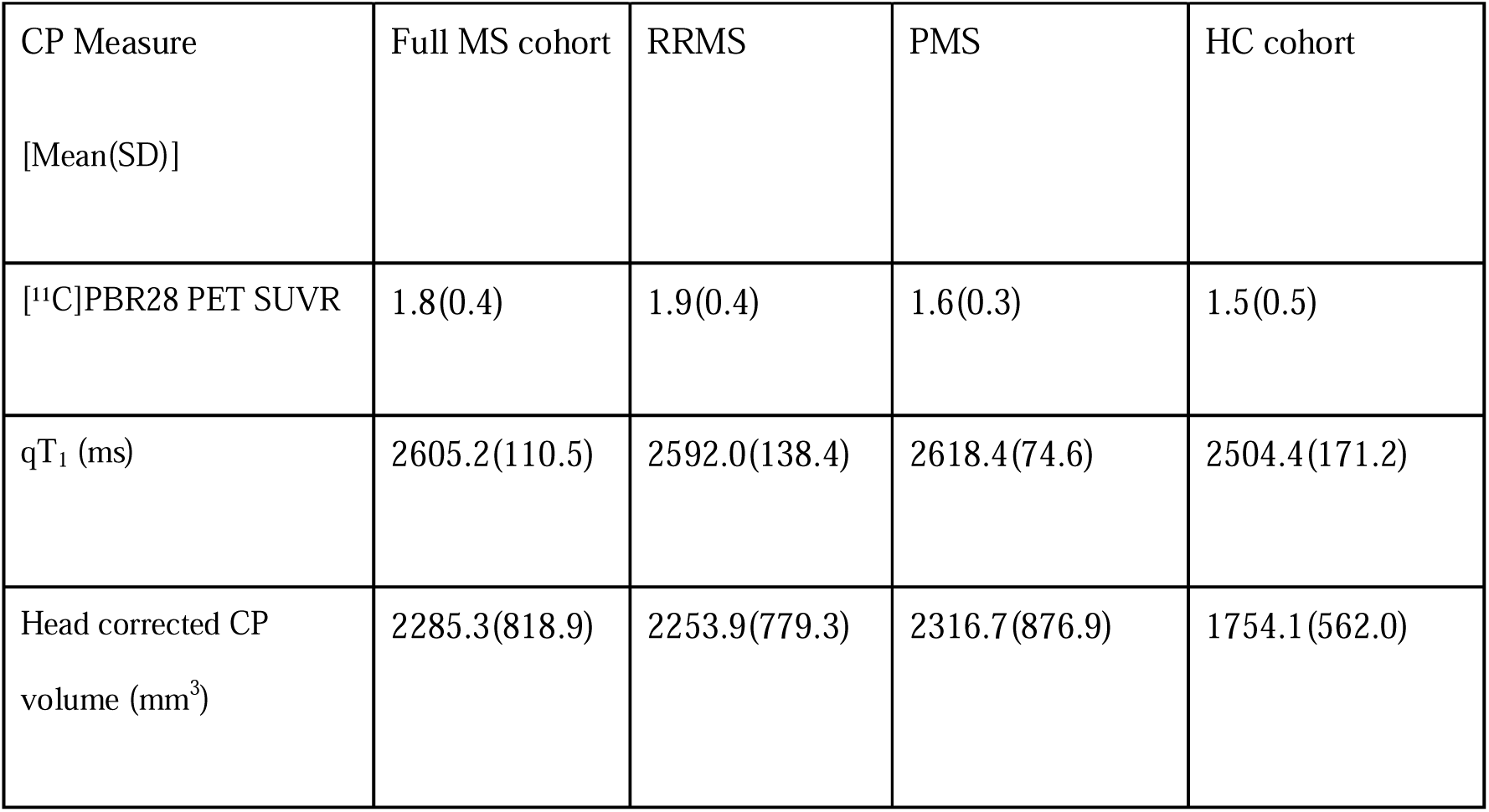
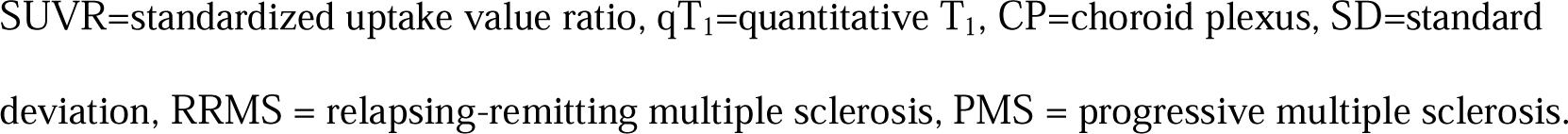
Summary of CP data derived from qT1 maps and SUVR maps for all participant groups.

| CP Measure<br>[Mean(SD)] | Full MS cohort | RRMS | PMS | HC cohort |
| --- | --- | --- | --- | --- |
| [$^{11}\text{C}$]PBR28 PET SUVR | 1.8(0.4) | 1.9(0.4) | 1.6(0.3) | 1.5(0.5) |
| $qT_1$ (ms) | 2605.2(110.5) | 2592.0(138.4) | 2618.4(74.6) | 2504.4(171.2) |
| Head corrected CP<br>volume ($\text{mm}^3$) | 2285.3(818.9) | 2253.9(779.3) | 2316.7(876.9) | 1754.1(562.0) |

SUVR=standardized uptake value ratio, $qT_1$ =quantitative $T_1$ , CP=choroid plexus, SD=standard deviation, RRMS = relapsing-remitting multiple sclerosis, PMS = progressive multiple sclerosis.

Mean qT□ values of the CP were also significantly prolonged in the MS group (p<0.05; Figure 3B), indicating increased water content or expansion of the extracellular space. Consistent with previous literature (10, 12, 28), head-size corrected CP volume was significantly greater in pwMS compared to controls (p<0.01;Figure 3C). Furthermore, in the pwMS cohort, partial Spearman correlation showed a significant inverse qT□–SUVR association (ρ_partial = -0.45, *p*<0.01). To determine whether this effect was driven by a particular MS subtype, we examined associations within the RRMS and PMS groups. In RRMS, the inverse relationship was moderately strong and statistically significant (ρ_partial = -0.54, *p*<0.05), whereas for PMS participants, the correlation was weaker and not significant (ρ_partial = -0.37, *p*= 0.22). Contrastingly, in controls, there was no association (ρ_partial

= -0.32, p = 0.19). Figure 4 shows the corresponding scatter plots, including linear fits and annotations of the partial Spearman correlation coefficients and p-values.

**Figure 4:**
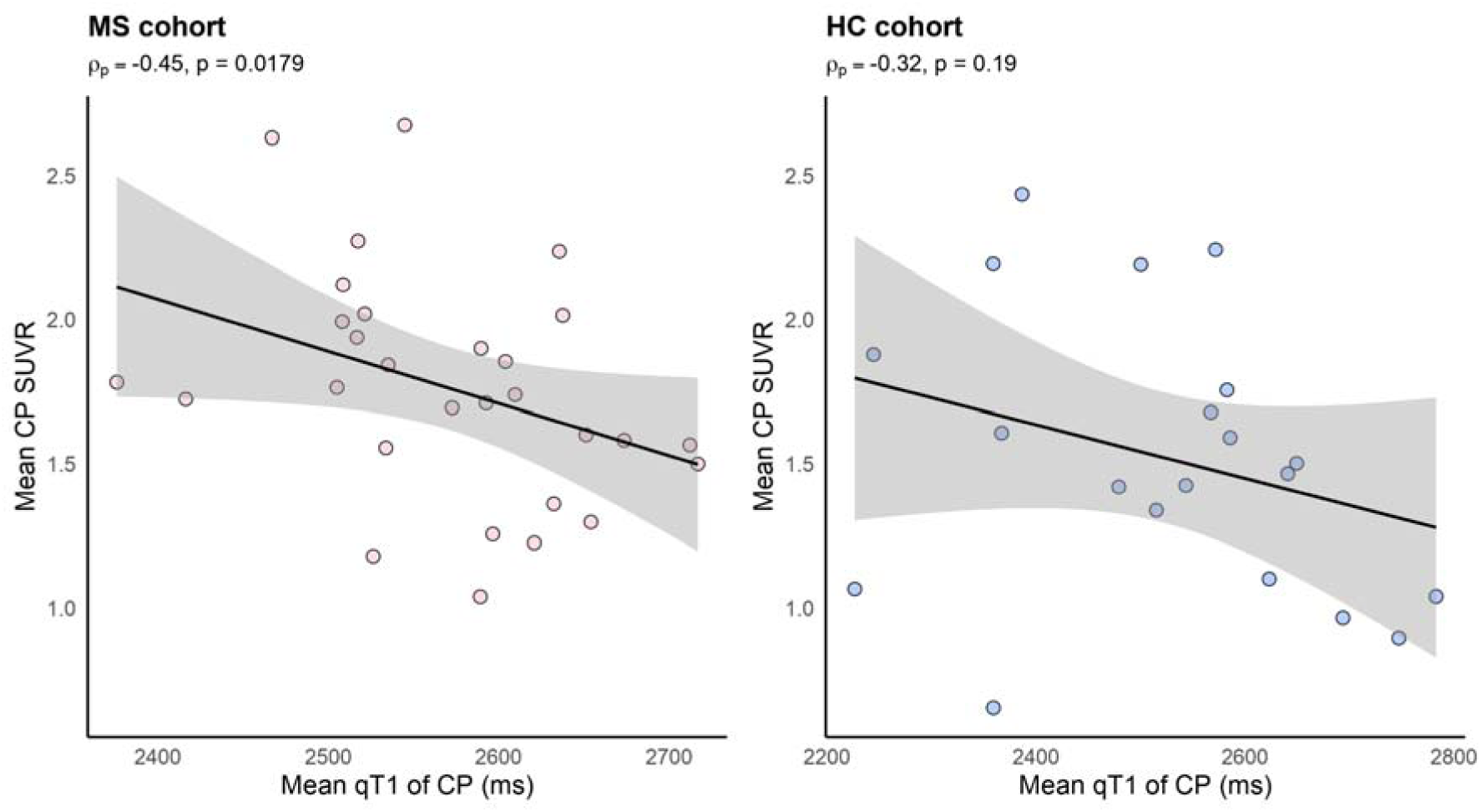
Scatter plots showing mean qT_₁_ of CP versus CP [¹¹C]PBR28 SUVR. Partial Spearman correlation analyses adjusted for age are shown separately for MS participants and healthy controls (HCs). A significant inverse association was observed in the pwMS cohort (ρ_partial = –0.47, *p*<0.01), while no significant association was found in HCs (ρ_partial = –0.34, *p*=0.14). Linear fits are displayed with 95% confidence intervals.

As demonstrated in Figure 5, CP volume showed a significant positive association with CP qT□ for all participants included in the study (*R*² = 0.275, *p*<0.001). Within the RRMS subgroup, this relationship was stronger (*R*² = 0.431, *p*<0.001). No significant association was observed between CP SUVR and CP volume in the pwMS cohort. Age and sex were not significant covariates in either of the aforementioned models and were, thus, omitted as covariates. No significant associations were observed between CP SUVRs (or CP qT_1_s) and disability scores, white□matter lesion burden or cortical lesion counts.

**Figure 5:**
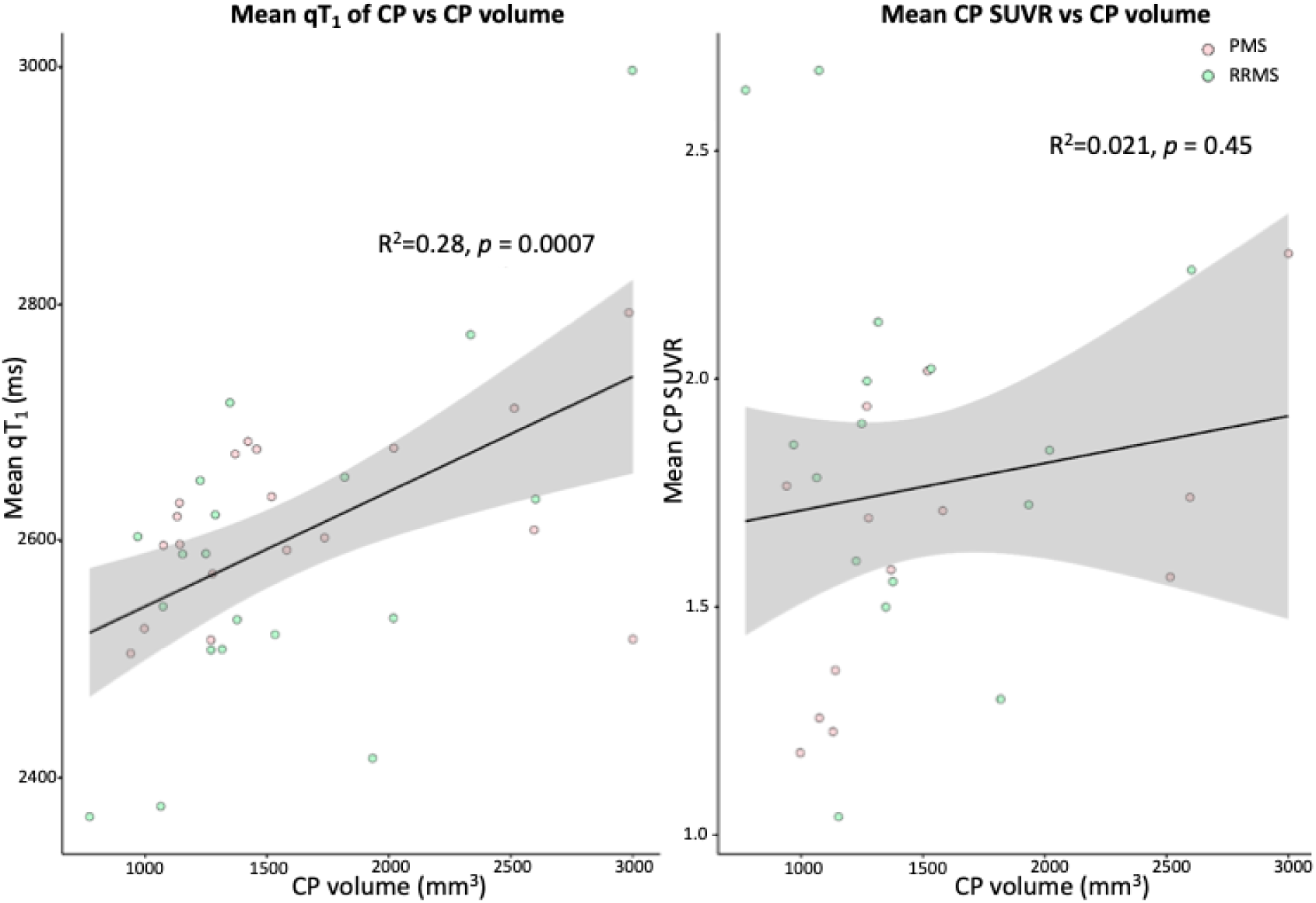
Linear regression models showing associations of CP [¹¹C]PBR28 SUVR and CP qT_₁_ with CP volume. CP volume showed a significant positive association with CP qT_₁_ (*r*=0.52, *p*<0.001). No significant associations were seen between mean CP [¹¹C]PBR28 SUVR and CP volume.

## Discussion

In this multimodal imaging study, we combined ultra–high-field MRI-based, quantitative TL mapping with high-resolution [¹¹C]PBR28 PET to investigate CP alterations in MS. We found that CP SUVR, a marker of TSPO expression, was higher in pwMS than in healthy controls. The observed elevations in CP SUVR were driven by the RRMS subgroup. This pattern is consistent with heightened innate immune activity at the blood–CSF interface that has previously been reported in relapsing MS (10). It may partially reflect recent relapses or MRI activity that was observed in 36.8% of pwMS enrolled in our study. Our findings are also aligned with prior reports (8, 10), providing further *in vivo* evidence that the CP is a site of heightened innate immune activity in MS.

The elevation in [¹¹C]PBR28 PET SUVR in the CP of pwMS provides evidence of increased TSPO availability, a marker commonly associated with microglial activation and/or reactive astrocytes (29). Prior PET imaging studies have also demonstrated more widespread TSPO upregulation in the MS brain, particularly in active lesions and periventricular regions, suggesting that innate immune activation is related to disease activity (30-32). Histopathological studies corroborate findings of higher activated microglial density in MS brain tissue and implicate the CP as a site of immune surveillance and activation. It has also been demonstrated that the CP in MS harbors activated phagocytic cells, including epiplexus macrophages and stromal monocytes, which express markers such as HLA-DR and CD68 and are capable of cytokine and chemokine release (5). These innate immune cell types may contribute to the increased TSPO PET signal observed in pwMS compared to controls (33).

In qMRI analyses, we observed CP qT□ was prolonged in MS, providing *in vivo* evidence of disease-related microstructural alterations within the plexus. Since qT□ is highly sensitive to tissue water content and integrity of the microstructural environment (34), increased qT□ likely reflects an expansion of the extracellular or interstitial compartment in the stroma, consistent with stromal remodelling within the CP (35). This interpretation is further supported by our findings of higher CP volumes in MS, consistent with previous reports (10, 12, 28, 36). It is also supported by the strong positive correlation between CP qT□ and CP volume. Expansion of the interstitial fluid compartment within the CP stroma has been powerfully modelled in Experimental Autoimmune Encephalomyelitis (EAE) where massive ultrastructural changes alongside upregulation of adhesion molecules like VCAM-1 and ICAM-1 on CP epithelial cells reflect barrier dysfunction (4). An underlying molecular mechanism, as elucidated by a recent single-cell transcriptomics study, suggests that stromal remodelling is not a passive byproduct of inflammation, but an active, orchestrated process directed by the CP epithelium itself in response to peripheral signals (37, 38). In this paradigm, peripheral inflammatory triggers activate NF-κB signaling within CP epithelial cells, driving the transcriptional upregulation of key adhesion molecules (e.g., VCAM-1) along with an array of cytokines and chemokines. This active epithelial–stromal crosstalk promotes the infiltration of monocytes and T cells, which facilitates the release of toxic factors, such as pro-inflammatory cytokines and reactive oxygen species, directly into the CSF (5). Histopathological findings support this model, showing that inflammatory epithelial cells recruit and activate macrophages, which adhere to the CSF-facing epithelium, amplify local pro-inflammatory signaling, and contribute to barrier remodelling through tight junction repair (39).

While our group-wise analyses showed elevated CP TSPO-PET SUVR and prolonged qT_1_ of CP in pwMS compared to controls, we observed a moderate inverse association between these two independent imaging metrics within the MS group. The inverse correlation was primarily driven by the RRMS participants. The absence of such an association in healthy controls likely suggests a disease-specific, dynamic relationship between neuroinflammation and stromal expansion at the blood-CSF barrier. The PMS subgroup, which mostly consisted of older participants with longer disease duration, displayed a lack of a clear association between CP SUVR and CP qT_1_. This could, potentially, be explained by chronic, heterogeneous remodelling where fibrosis and vascular alterations that possibly decouple PET uptake from tissue microstructure (40, 41). That hypothesis is consistent with prior observations where, in some PMS cases, the inflammatory response approximates that of age-matched controls (41). Our findings in the RRMS cohort could biologically be explained by two mechanisms, one occurring at the epithelium (stromal remodelling) and the other at the fenestrated vessels of the BCSFB (vascular remodelling). Stromal remodelling due to dysfunction of the tight junctions within the CP epithelium could cause vasogenic edema. This would expand the extracellular water pool, prolonging qT□, while diluting TSPO-expressing cells within a voxel. Such a process would lower SUVR, despite ongoing inflammation, creating an ‘inflammatory dilution effect’. Altered fluid dynamics at the BCSF interface due to vascular remodeling could also reverse fluid flux from CSF into the stroma and effectively wash out both inflammatory mediators and TSPO signal near the epithelium.

There are several limitations to this study that should be considered. The resolution of PET imaging is limited, even when applying our high-resolution PET tomograph system. This is relevant as the CP is a small and irregularly-shaped structure. Although PVC was applied, using the Labbé + RBV algorithm, to all PET images, such images remain susceptible to inaccuracies when measuring SUVR in very small structures like the CP. This is especially true after image upsampling, as spill-in from CSF voxels may still confound quantification (42, 43). Additionally, TSPO is expressed not only by microglia but also by endothelial cells and reactive astrocytes. The [^11^C]PBR28 TSPO PET SUVR signal measured in our study may have been influenced by these cell types and this could have impacted the PET-derived interpretation of neuroinflammatory activity (33, 44, 45). Moreover, the sample size (n=34) for our study was modest and the cross-sectional design prevented assessment of longitudinal dynamics of CP imaging markers. Future work should investigate whether modern DMTs, such as ocrelizumab or fingolimod, have downstream effects on longitudinal CP inflammation, as existing studies have largely focused on time-dependent changes in gray matter and white matter, rather than CP-specific changes (46, 47).

Despite the aforementioned limitations, our findings of *in vivo* CP alterations in MS, as assessed by multimodal imaging markers, are consistent with experimental models and histopathological reports. They highlight the CP as a potentially important contributor to disease pathology. Future studies should also explore the role of oxidative damage in this framework, using approaches like quantitative susceptibility mapping or magnetic resonance spectroscopy. Furthermore, longitudinal imaging and molecular profiling could help understand the interplay between BCSF barrier integrity, microglial activation, and neuroinflammatory signalling during MS progression.

## Conclusion

Our findings provide *in vivo* evidence that the CP may undergo stromal/vascular remodelling and innate immune activation in MS and that these features may be detectable using ultra–high-field qT□ mapping and high-resolution [¹¹C]PBR28 PET. We posit that a dynamic remodelling process takes place at the blood–CSF barrier, rather than CP abnormalities being a passive byproduct of disease. Overall, our results position the CP as both a sensitive biomarker of neuroinflammatory activity and a potential therapeutic target in MS, warranting longitudinal imaging studies to define the role of the CP in disease progression and treatment response.

## Authorship Contribution statement

**Sneha Senthil:** Writing – review and editing, Writing – original draft, Visualization, Validation, Software, Methodology, Investigation, Formal Analysis, Data curation, Conceptualization. **Risavarshni Thevakumaran**: Writing – review and editing, Visualization, Validation, Resources. **Stephan Blinder:** Writing – review and editing, Validation, Software, Methodology, Resources, Formal Analysis. **Alexey Kostikov:** Writing – review and editing, Validation, Methodology, Resources, Funding acquisition. **Rozalia Arnaoutelis:** Writing – review and editing, Data curation, Resources. **Pedro Rosa-Neto**: Writing – review and editing, Resources, Funding acquisition. **G. R. Wayne Moore**: Writing – review and editing, Data curation, Resources. **Douglas L. Arnold:** Writing – review and editing, Resources, Funding acquisition. **Sridar Narayanan:** Writing – review and editing, Supervision, Resources, Data curation, Conceptualization, Funding acquisition. **David A. Rudko:** Writing – review and editing, Writing – original draft, Supervision, Resources, Software, Project administration, Methodology, Investigation, Formal Analysis, Data curation, Conceptualization, Funding acquisition.

## Acknowledgements

This study was supported in part by the United States Department of Defense, Multiple Sclerosis Research Program, Investigator-Initiated Research Award (Award No. W81XWH19-1-0486) (DAR). SS thanks the Fonds de Recherche Québec – Santé for personal support (scholarship). RT was supported by the Canada Graduate Scholarships Doctoral Award (CGS-D). The authors are grateful to all the participants who made this study possible. We gratefully thank the personnel of the PET radiochemistry unit at the McConnell Brain Imaging Centre, notably Robert Hopewell, Karen Ross, Monica Somoila, I-Huang Tsai and Dean Jolly. We also thank our excellent MRI technologists Ronaldo Lopez, David Costa and Soheil Mollamohseni Quchani and PET technologists Chris Hsiao and Catherine Saleh for their assistance.

## Data availability statement

The data that support the findings of this study are not publicly available due to concerns surrounding patient confidentiality but are available from the corresponding author to qualified investigators on reasonable request.

